# Characterizing the clinical adoption of medical AI through U.S. insurance claims

**DOI:** 10.1101/2023.08.26.23294629

**Authors:** Kevin Wu, Eric Wu, Brandon Theodorou, Weixin Liang, Christina Mack, Lucas Glass, Jimeng Sun, James Zou

## Abstract

There are now over 500 medical AI devices that are approved by the U.S. FDA. However, little is known about where and how often these devices are actually used after regulatory approval. In this paper, we systematically quantify the adoption and usage of medical AI in the U.S. by tracking Current Procedural Terminology (CPT) codes explicitly created for medical AI. CPT codes are widely used for documenting billing and payment for medical procedures, providing a measure of device utilization across different clinical settings. We examine a comprehensive nationwide claims database of 16 billion CPT claims between 1/1/2015 to 6/12023 to analyze the prevalence of medical AI based on submitted claims. Our results indicate that medical AI adoption is still nascent, with most usage driven by a handful of leading devices. For example, only AI devices used for assessing coronary artery disease and for diagnosing diabetic retinopathy have accumulated more than 10,000 CPT claims. Furthermore, medical AI usage is moderately over-represented in higher-income zip codes and metropolitan areas. Our study sheds light on the current landscape of medical AI adoption and usage in the U.S., underscoring the need to further investigate barriers and incentives to promote equitable access and broader integration of AI technologies in healthcare.

## Introduction

As artificial intelligence (AI) has rapidly progressed in recent years, significant investments have been devoted to developing and commercializing AI in medicine. As of 2023, over 500 medical AI devices have undergone U.S. FDA evaluation and received approval across areas such as radiology, neurology, and pathology^1^. During an FDA submission, device manufacturers are required to report evidence of the efficacy and safety of their products, providing crucial insight into how AI algorithms are evaluated before being used on patients^2^. However, after approval, companies rarely share where and when their products are used. As such, despite the proliferation of medical AI approvals, little is known about their real-world usage.

The usage and adoption patterns of medical AI can significantly impact their clinical impact. First, the performances of AI algorithms are notoriously susceptible to changes in healthcare settings and fluctuate during deployment^3,4^. For instance, despite initial studies indicating up to a 20% improvement in detection rates, computer-aided detection (CAD) products for mammography approved in the early 2000s have been found to provide no tangible benefits to women^5^. This discrepancy has been attributed to adoption and usage factors such as changes in clinician interaction with the software and the transition from film to digital mammograms^6^. Consequently, while AI medical devices may demonstrate strong performance under specific evaluation conditions, variations in real-world applications can yield drastically different outcomes. Second, the impact of medical AI devices is mediated by economic forces. After FDA approval, companies need to find sustainable revenue streams for the promises of AI-driven healthcare to be realized. Different reimbursement approaches can affect how often and on whom these devices are used, and it is still unclear which model is optimal for the new AI devices^7,8^. Studying the empirical usage of medical AI devices is a crucial step in characterizing the landscape of medical innovations and can provide a more holistic view of the translational pipeline from algorithm to patient.

Recently, Current Procedural Terminology (CPT) codes have been created specifically for medical AI devices^7,8^. CPT codes are designated by the U.S. Department of Health and Human Services under the Health Insurance Portability and Accountability Act (HIPAA) as a national coding set for physicians and other healthcare professional services and procedures to be used by the Centers for Medicare & Medicaid Services (CMS)^9^. The codes are regularly created, updated, modified by the American Medical Association (AMA), and are the most widely accepted medical nomenclature under public and private health insurance programs^9^. Healthcare providers use these codes to generate itemized bills detailing the specific services delivered to a patient during a medical encounter. Subsequently, these bills are submitted to insurance companies, who use the coded information to determine the appropriate reimbursement for the services rendered. As such, CPT codes play a crucial role in ensuring the accuracy and uniformity of medical billing, as well as promoting accountability and transparency within the healthcare system.

CMS also provides coverage for medical AI through New Technology Add-On Payment (NTAP) which is specifically designed to encourage healthcare providers to adopt new technologies^7^. However, NTAP specifically focuses on inpatient payments, whereas CPT codes apply to both inpatient and outpatient settings^10,11^. In this paper, we focus on CPT codes as they are most widely adopted and standardized across both public and private insurance programs^9^, whereas NTAP is specifically used within Medicare^11^, presenting only a partial view of national AI usage. Additionally, due to its extensive and long-term adoption by healthcare payers, CPT is also an informative resource for comparing baseline usage rates of non-AI devices.

While an increasing number of CPT codes have been made available for medical AI devices, these codes are generally spread across various medical domains and reserved for medical coders and insurance companies. As such, there currently does not exist a single database of AI-related CPT codes or a systematic analysis of their usage. In this paper, we identify and organize a comprehensive list of CPT codes that apply to medical AI devices. We analyze the usage of these codes on a large national claims database and present their temporal and geographical trends.

### Related works

Previous analyses have focused on translational roadblocks for medical AI stemming from model evaluation, ethics, and reporting^2,12^. Specific studies have shown how AI algorithms can perform worse in clinical practice despite promising retrospective evaluations^13,14^. A variety of studies have analyzed the emergence of reimbursement mechanisms for medical AI products. For example, researchers have highlighted Viz.AI’s NTAP payment model and its potential impact on stroke care, as well as the economic challenges of adopting LumineticsCore from a cost-benefit perspective^11,15^. Current payment models for AI have been previously analyzed along with examples of reimbursable AI devices^7^. More specifically, a recent study has proposed a framework for analytically determining the value and cost of each unique AI service in order to encourage ethical and optimal deployment^8^.

While our work is the first to analyze AI usage through CPT codes, several studies have analyzed geographical distributions present in AI development. For example, researchers have analyzed PubMed for the training datasets used in various medical AI algorithms and found that the data are disproportionately located in California, Massachusetts, and New York^16^. Datasets used in AI skin cancer diagnosis have also been exclusively found to be from Europe, North America, and Oceania^17^. The usage of CPT codes for digital health technologies like remote physiologic monitoring, eConsults, and eVisits have also been systematically studied by reporting the total number of claims in Medicare data^18^. Our work focuses specifically on the subset of digital health relevant to artificial intelligence and machine learning.

## Methods

Our analysis consists of two main parts: the organization of medical AI CPT codes and the analysis of their usage. First, to find CPT codes used for medical AI devices, we use a combination of official sources, web resources, and insurance company policies. Second, we searched a large national claims database to quantify the usage of each code.

### Collecting CPT Codes for Medical AI

#### Official AMA sources

The AMA develops CPT codes and is responsible for the development of new billing codes for medical AI products. The CPT Editorial Panel has issued guidance for classifying AI applications (Appendix S), which includes assistive, augmentative, and autonomous work^19^, but only a few examples of AI codes are referenced. For a comprehensive list of new CPT codes, we processed the AMA’s list of Category III codes (accessed 3/1/23 ^20^), which are a set of temporary codes assigned to emerging technologies, services, and procedures^21^. While these codes are billed like all other codes, Category III codes are intended to be used primarily for data collection to substantiate widespread usage before granting reimbursement. After five years, they are re-evaluated and replaced with a Category I code if deemed qualified. We analyzed each of the AMA’s Category III codes (long descriptors) for the terms *artificial intelligence* and *machine learning* and their variants. Next, for Category I and II codes, we perform a comprehensive search using Codify by AAPC, a search engine for CPT codes^22^.

CPT code long descriptors provide limited information on the underlying technology behind the procedure and the product name. As such, we complement the CPT code descriptions with details provided by insurance companies in policy documents. Such documents provide detailed descriptions of a given procedure, as well as any medical evidence that might support the case for its reimbursement. Additionally, the policies often reference specific product names that the CPT codes refer to. We analyzed the policies of Premera, Amerigroup, and Blue Cross and noted products that were referred to as artificial intelligence or machine learning devices^23–25^.

#### Determining AI devices

We determine whether each candidate CPT code bills for an AI medical device if either of the following criteria is met: (1) the device manufacturer makes explicit marketing claims that its product uses AI and/or machine learning (ML), or (2) a third-party (e.g. insurance company, news publication) refers to the product as powered by AI and/or ML. Additionally, we exclude CPT codes which are also used for billing non-AI devices, as this dilutes the number of AI-specific occurrences. For example, recently, AI has been integrated into a continuous glucose monitoring device, but other non-AI devices are billed under the same code. Another example includes mammography with computer-aided detection (CAD), which is largely dominated by traditional CAD and should be differentiated from modern CAD products^26^. Next, several CPT codes exist for machine learning-based proprietary lab tests (identified with the letter ’U’), but are excluded from this study since they are designed for specific laboratories and are typically not reimbursed by third-party payers. Finally, to focus our analysis on the usage of recently developed AI, we only include CPT codes developed after 2015.

#### Grouping CPT Codes

Multiple CPT codes may be related to the same underlying medical procedure but describe different aspects of the procedure. For example, both 0648T and 0649T are used to report quantitative magnetic resonance analysis of tissue composition, but 0649T is used when a diagnostic MRI is also completed, while 0648T is used when it is not. In our analysis, we organize codes that refer to the same underlying medical AI procedure into a CPT code group. To this end, we compute the sum total of all codes in that code group when reporting the number of claims for each procedure.

### Claims Data

#### IQVIA PharMetrics® Plus

We use the IQVIA PharMetrics® Plus for MedTech dataset, a longitudinal health plan database of medical and pharmacy claims^27^. The dataset consists of more than 210 million unique U.S. enrollees and is comprised largely of commercial health plans. The data are compliant with the Health Insurance Portability and Accountability Act (HIPPA) and are representative of the commercially insured US national population for patients under 65 years of age^28^. The IQVIA dataset is commonly used for analyses of medical trends in areas like infectious diseases^29–33^, cardiology^34^, dermatology^35^, pulmonology^36^, oncology^37^, and neurology^38–40^. The unit measurement we use in our analysis is a medical claim that uses a CPT code associated with a medical AI procedure. We analyze usage in all 50 U.S. states from 01/01/2015 to 06/01/2023; the dataset consists of 16 billion claims in total. As a point of reference, CMS reports that there are a total of 5 billion claims processed in the US per year ^41^, which suggests that our dataset has around 40% coverage of all US claims.

#### Finding Associated Device Names and FDA Approvals

To provide the commercial context for each CPT code, we also locate specific device names associated with each AI CPT code by searching through insurance policies as well as company websites. While we were able to locate at least one product for each procedure, the list may not be comprehensive if not indicated by the company or a third-party source. For the top products we found, we also located their corresponding FDA approval (if applicable) to provide a timeline context for the overall translational pipeline for each product.

#### Geographic analysis

For each medical AI procedure, we aggregate all unique zip codes that contain an occurrence of at least one code. First, we search for each zip code’s median income and classify it as high-income if it exceeds $100,000, consistent with the IRS’s classification^42^. Next, we determine whether it is in a metropolitan area by referencing the U.S. Department of Agriculture’s Rural-Urban Commuting Area Codes (RUCA)^43^. Finally, we compute the percent of all unique zip codes that (1) have a high median income and (2) are metropolitan. We compare these rates to the rates found for all US zip codes, as well as unique zip codes found in a random sample of one million claims (across all CPT codes).

#### Insurance Pricing

In addition to CPT code billing frequencies, we collect public and private pricing information. First, when available, we look up Medicare pricing for each CPT code that has been made publicly available each year^44^. Second, we gather negotiated pricing rate data from Anthem Healthcare in California and New York, focusing specifically on in-network rates as of November 2022. This data is made available as part of the Transparency in Coverage (TIC) regulation, which was introduced by the Tri-Agencies (U.S. Departments of Health and Human Services, Labor, and Treasury) on 11/12/2020^45^. The regulation requires health plans to publish their negotiated rates for all items and services for commercial coverage, including in-network files, in machine-readable formats, with monthly updates starting from July 1, 2022. We utilize the November 2022 version of the in-network rate files, which are provided in the Centers for Medicare & Medicaid Services (CMS) defined JSON format.

## Results

### Billable Medical AI

Given our methodology, we find a total of 16 medical AI procedures billable under CPT codes. Several procedures can be reimbursed through multiple codes, comprising a total of 32 unique CPT codes that are associated with AI. These procedures are detailed in Table 1, alongside the total number of claims containing the codes, product name, and effective date of the codes. The procedures fall within a wide range of healthcare areas, such as cardiology, radiology, and ophthalmology, and have been created very recently, with 15 of 16 medical AI procedures created since 2021 (Figure 1). We find that only 4 out of 16 have more than 1,000 total claims. This is partially because the median age of a medical AI procedure is only about a year (374 days) old.

**Figure 1:**
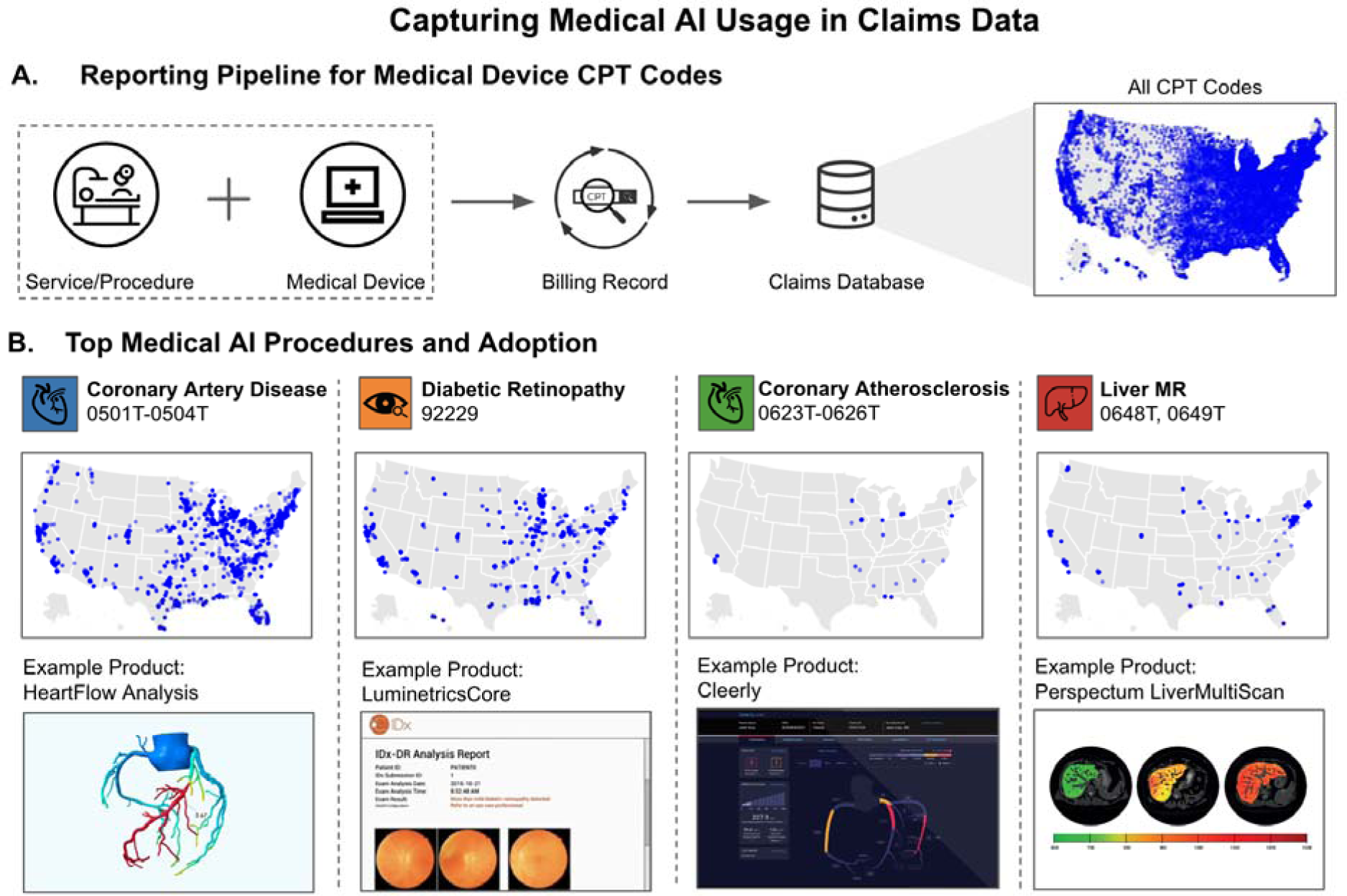
Capturing Medical AI Usage in Claims Data. (A) Medical device usage is captured through billing records and aggregated in our claims database. Each service/procedure that uses a medical device is associated with a CPT code that hospitals and medical practices report for billing purposes. On the right, we provide a map of the geographical distribution of zip codes for a random sample of 1 million claims (out of 16B in our dataset from 01/01/2015-06/01/2023) for comparison with AI CPT codes. Each blue dot represents a single unique zip code where the procedure was billed. (B) We provide details on the top four billable medical AI procedures through CPT codes. Under the procedure name, we list the CPT code(s) and a map of the geographical distribution of zip codes where they have been billed (cumulative over time). Finally, in the bottom row, we provide examples of billable products under each code and a product description image taken from marketing materials from the respective companies.

**Table 1:** Summary of AI CPT codes. A total of 16 medical AI procedures are presented alongside their corresponding CPT code(s). Each procedure is associated with an example commercial product that may be reimbursed through the codes. The effective date is the date on which the code is officially recognized by the American Medical Association (AMA) and can be used for billing and reimbursement purposes. The total claims listed are recent as of 06/01/2023.

| Total Claims | Medical AI Procedure | CPT Code(s) | Example Product Name | Effective Date |
| --- | --- | --- | --- | --- |
| 67306 | Coronary Artery Disease | 0501T-0504T | HeartFlow Analysis<sup>55</sup> | 06/01/2018 |
| 15097 | Diabetic Retinopathy | 92229 | LumineticsCore<sup>54</sup> | 01/01/2021 |
| 4459 | Coronary Atherosclerosis | 0623T-0626T | Cleery<sup>56</sup> | 01/01/2021 |
| 2428 | Liver MR | 0648T-0649T | Perspectum LiverMultiScan<sup>56</sup> | 01/01/2021 |
| 591 | Multi-Organ MRI | 0697T-0698T | Perspectum CoverScan<sup>57</sup> | 01/01/2022 |
| 552 | Breast Ultrasound | 0689T-0690T | Koios DS<sup>58</sup> | 01/01/2022 |
| 435 | ECG Cardiac Dysfunction | 0764T-0765T | Anumana<sup>59</sup> | 01/01/2023 |
| 331 | Cardiac Acoustic Waveform Recording | 0716T | CADScor<sup>61</sup> | 07/01/2022 |
| 237 | Quantitative MR Cholangiopancreatography | 0723T-0724T | Perspectum MRCP+<sup>59</sup> | 07/01/2022 |
| 67 | Epidural Infusion | 0777T | Compuflo<sup>60</sup> | 01/01/2023 |
| 4 | Quantitative CT Tissue Characterization | 0721T-0722T | Optellum Virtual Nodule Clinic<sup>61</sup> | 07/01/2022 |
| 1 | Autonomous Insulin Dosage | 0740T-0741T | d-Nav<sup>62</sup> | 01/01/2023 |
| 1 | CT Vertebral Fracture Assessment | 0691T | HealthVCF<sup>65</sup> | 01/01/2022 |
| 1 | Noninvasive Arterial Plaque Analysis | 0710T-0713T | ElucidVivo<sup>59</sup> | 01/01/2022 |
| 0 | Facial Phenotype Analysis | 0731T | Face2Gene<sup>56</sup> | 07/01/2022 |
| 0 | X-Ray Bone Density | 0749T | OsteoApp<sup>63</sup> | 01/01/2023 |

### Growth Patterns of Medical AI

We find that the overall utilization of medical AI products is still limited and focused on a few leading procedures. However, utilization has generally increased exponentially for each medical AI procedure (Figure 2). The procedure with the most AI usage is coronary artery disease (n=67306, effective 01/01/2018). The associated CPT codes can be used to reimburse products like HeartFlow FFRCT, a medical device that uses computed tomography (CT) scans to create a 3D model of the coronary arteries. The model is then used to calculate the fractional flow reserve (FFR), which is a measure of how well blood flows through the arteries. Among other functions, FFRct can be used to diagnose coronary artery disease and assess the severity of the disease^46^. HeartFlow FFRCT was given its first FDA approval in 2019 and has had two subsequent updates since^47^. In November of 2021, CMS set the national payment rate of the device at $930.34 for an office-based setting^48^.

**Figure 2:**
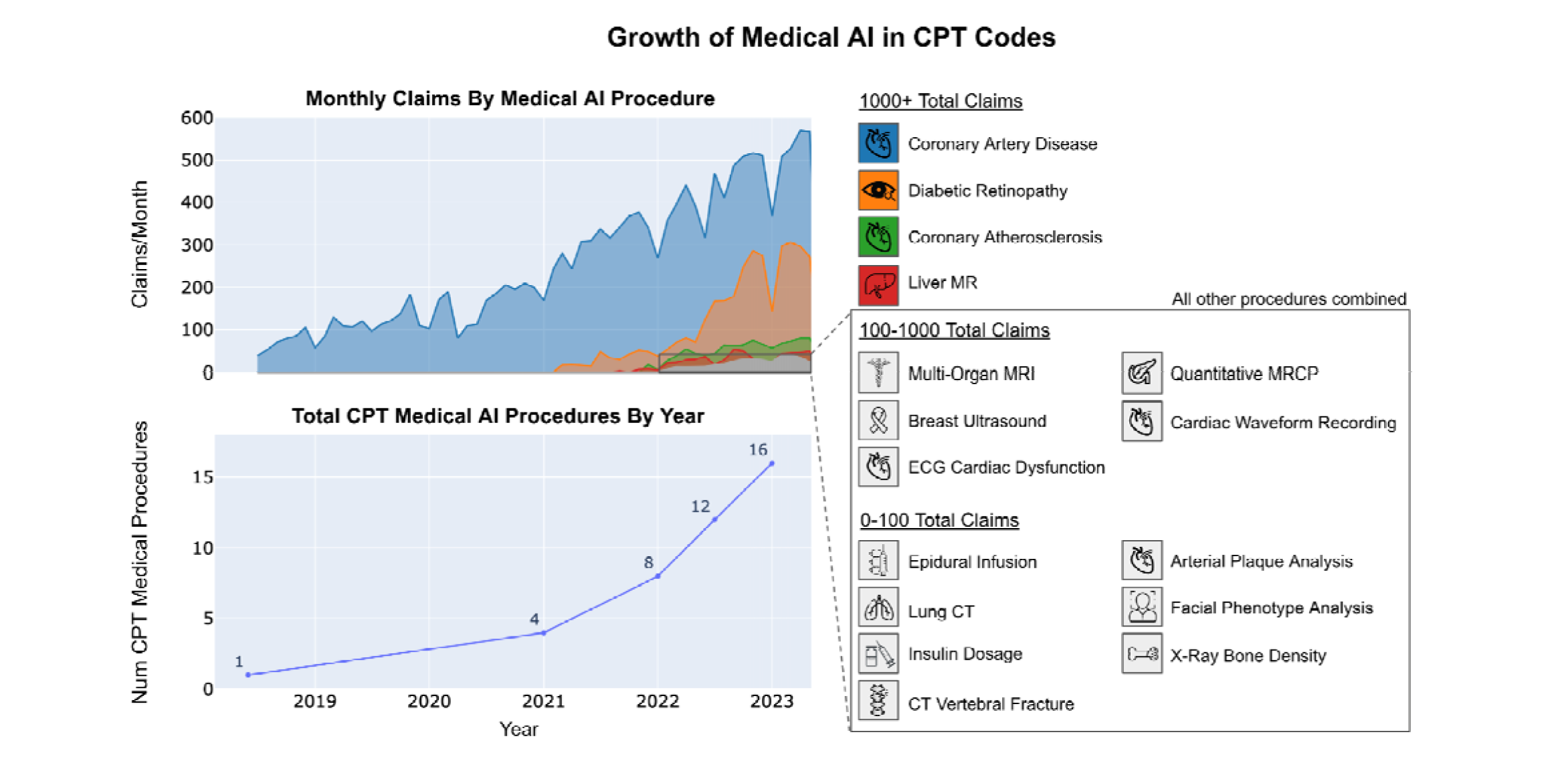
Growth of Medical AI in CPT Codes. (Top) The number of claims per month of each medical AI procedure between the dates of 01/01/2018 to 06/01/2023. The top four procedures by total claims are presented in colors, while the remaining 12 are grouped and added together into an "Other" group in gray. On the right-hand side, we provide a legend for each of the medical AI procedures. These procedures are further grouped by their usage tiers (0-100, 100-1000, and 1000+ total claims). All procedures in the "Other" category are contained in the callout box on the bottom right. (Bottom) We present the cumulative number of CPT AI medical procedures available each year from 2018-2023.

Diabetic retinopathy medical AI has also grown exponentially in usage (n=15097, effective 01/01/2021). The first FDA approval in this category was given on 1/12/2018 to LumineticsCore^49^, an AI diagnostic system that autonomously diagnoses patients for diabetic retinopathy (including macular edema)^50^. It is indicated for use by healthcare providers to automatically detect more than mild diabetic retinopathy (mtmDR) in adults diagnosed with diabetes who have not been previously diagnosed with diabetic retinopathy^49^. The product takes images of the back of the eye, analyzes them, and provides a diagnosis. If more than a mild case is detected, the patient is referred to a specialist^51^. In 2021, the national payment rate set by CMS for CPT code 92229 was $45.36^52^.

We also find exponential growth at a smaller scale occurring in medical AI for coronary atherosclerosis and liver MR. Cleery’s Coronary Computer Tomography Angiography (CCTA) algorithm (n=4459, effective 01/01/2021) received its first FDA approval on 10/9/2019 and aims to identify atherosclerosis, the plaque buildup in the arteries of the heart, as well as vascular morphology features for all identified arteries in the CCTA data^53^. While pricing for this code is not given through CMS, we find it has a median negotiated rate of $371.55 in Anthem’s CA and NY pricing data. Perspectum’s LiverMultiScan (n=2428, effective 01/01/2021) is a non-invasive diagnostic technology for evaluating liver diseases present in multiparametric MRI by quantifying liver tissue^54^. Receiving its FDA approval on 09/06/2017, it provides a number of quantification tools, such as Region of Interest (ROI) placements, to be used for the assessment of regions of an image to aid in the diagnosis of liver disorders^47^. The associated CPT code, 0648T, does not have a national payment rate through CMS, given its status as a Category III code. However, in Anthem’s CA and NY pricing data, we find that it reimburses for a median negotiated rate of $692.91.

Finally, we also observe that several procedures have had only nominal or zero usage. CT Vertebral Fracture Assessment and Noninvasive Arterial Plaque Analysis only had a single occurrence in our CPT database since 01/01/2022, and procedures (Facial Phenotype Analysis and X-Ray Bone Density) did not have any occurrences in our database.

### Characteristics of Deployed Zip Codes

Next, we analyze the unique zip codes where medical AI procedures are deployed. We find that 32% of zip codes where AI are deployed are high-income (>$100k median household income), which is significantly higher than non-AI claims (17%, p<0.001), as well as the US general population average (10%, p<0.001). An average of 89% of the zip codes for AI are metropolitan, which is much higher than the US average (41%, p<0.001), and marginally higher compared to the random sample of non-AI claims (87%, p=0.002) (Figure 3). Additionally, we provide a map of the geographical distribution of claims for the top 4 medical AI procedures for each year of their availability and find that usage is generally well distributed across coasts and regions of the US (Supp. Figure 1). Additionally, we list out the top 10 zip codes and city/state for each of the top 4 medical AI procedures in our analysis (Supp. Table 1).

**Figure 3:**
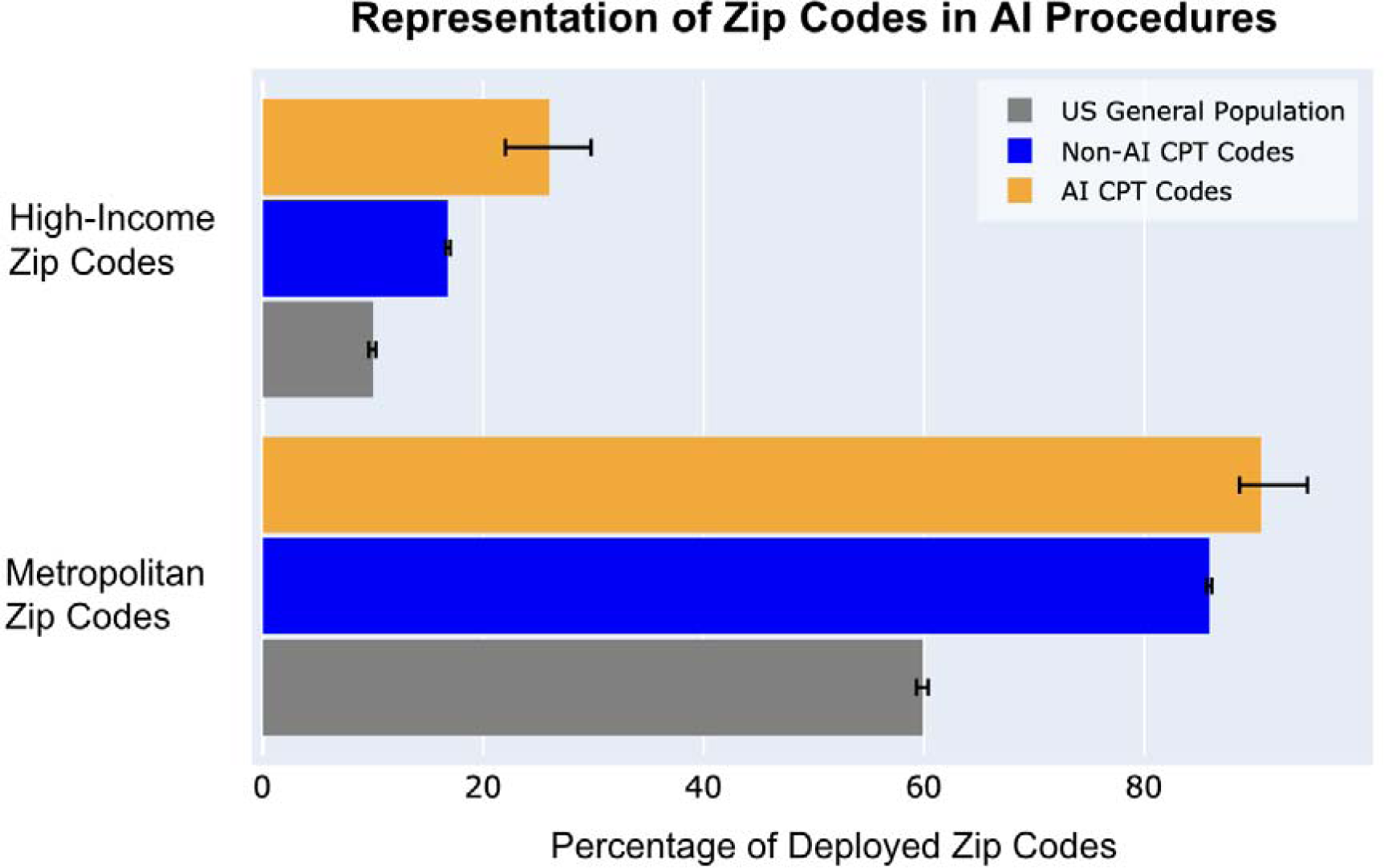
Representation of Zip Codes in AI Procedures. All unique zip codes are categorized by income level and metropolitan area. The top row shows the percentage of all unique zip codes that have a median income above $100K for AI devices, all medical devices (including non-AI), and the general US population. The bottom row shows the percentage of metropolitan zip codes across the same three categories. The black bars represent 95% confidence intervals based on bootstrapped samples.

## Discussion

Our study finds that the commercialization of FDA-approved AI products is still nascent but growing, with over 50% of CPT codes effective since 2022. However, only a handful of these devices have reached substantial market adoption, suggesting that the medical AI landscape is still in its early stages. Such usage patterns underscore key themes regarding the deployment of AI in medicine, including clinical implementation challenges, payment, and equal access.

Successful clinical adoption of medical AI involves overcoming key implementation barriers. First, the addition of AI may require significant changes to the clinical workflow. For example, studies have detailed how the success of a diabetic retinopathy detection algorithm is mediated by deployment factors like patient consent, internet speed and connectivity, and poor lighting conditions^64,65^. Another study finds that the added benefit of an AI algorithm in pathology depends on the pathologist’s interaction with the algorithm’s outputs^66^. Moreover, the value of an AI algorithm is to clinical practices^67^. For instance, researchers have argued that clinics that use diabetic retinopathy algorithms may operate at a deficit for every patient evaluated and propose modifications to the existing payment structure to encourage adoption^15^. On the other hand, patients may be incentivized to visit practices that provide state-of-the-art technologies. Medical AI devices need to have a clear value proposition to healthcare providers to achieve widespread adoption, but the value of AI is multifaceted and context-dependent^68,69^.

In particular, Medicare pricing for medical AI can provide insight into how AI is currently valued. The reimbursement amounts for CPT codes are determined based on three factors: physician work, practice expense, and malpractice cost^70^. For a given code, each factor is associated with a relative value unit (RVU) that is adjusted to account for differences between procedures. For example, a higher RVU for physician work means the procedure involves more physician time and/or expertise. A key value proposition of medical AI devices is their ability to reduce or remove the work burden of physicians. We find this reflected in the pricing for CPT code 92229 (diabetic retinopathy) in the CMS fee schedule. Despite having a relative value of 0 for physician work, the practice expense relative value (peRVU) for this code is 1.34, which is higher than its non-AI counterpart (CPT code 92228, peRVU=0.53)^71^. This difference illustrates how the pricing of AI devices shifts some of the value typically assigned to physicians toward the costs of purchasing and operating the device itself.

While we find an increasing rate of utilization of medical AI for several market leaders, we also note disparities in the adoption of these technologies across different healthcare settings. Our findings indicate that, on average, AI devices are deployed in wealthier zip codes at a higher rate than a random sample of claims for all CPT codes, especially in the initial stages of deployment. Such disparities could be due to various factors, such as the cost of the devices and the financial resources required to implement and maintain these technologies. Additionally, healthcare providers in wealthier areas may be more inclined to invest in cutting-edge technologies to attract patients and maintain a competitive edge. Policymakers and stakeholders should implement strategies that promote equitable access to medical AI technologies to prevent further widening of existing healthcare disparities.

Our analysis of medical AI usage has several limitations. First, while our dataset of 16 billion claims (IQVIA PharMetrics® Plus) is representative of the US patient population under 65, it does not capture all medical claims. As such, the number of claims reported in our work only represents a fraction of total usage and should mainly be interpreted through its relative magnitude over time. Second, our analysis focuses specifically on CPT codes, which do not capture all potential types of AI usage. For example, products such as Viz.ai’s LVO detection algorithm are reimbursable under Medicare’s NTAP program, but we do not capture such usage in our study. Additionally, medical AI usage in clinical pilot studies that are not reimbursed will not appear in large national databases. Our analysis also does not capture the usage of medical AI devices that are billed under non-AI-specific CPT codes. For example, CPT code 77066 is used for computer-assisted detection (CAD) for mammograms but does not differentiate the usage of current deep learning-based approaches from older traditional models from the 1990s. As such, while new models for mammography are developed and approved by the FDA, their usage can not be cleanly identified in claims data.

The usage and adoption of medical AI are the product of a complex ecosystem involving AI developers, healthcare providers, payers, and patients. While the last few years have seen rapid growth in the capabilities of AI, careful consideration of forces beyond algorithmic development is required for AI models to have a meaningful clinical impact. As such, monitoring the usage and clinical adoption of medical AI is key to ensuring that these new technologies fulfill the promise of improving the quality of healthcare for broad patient populations.

## Data Availability

The IQVIA Pharmetrics Plus claims dataset utilized in this research is available for licensing or for research through IQVIA. The insurance pricing data employed in this study is publicly accessible as part of the Transparency in Coverage (TIC) regulation and can be obtained directly through CMS at https://www.cms.gov/healthplan-price-transparency.

## Data Availability

All data produced in the present study are available upon reasonable request to the authors

## Acknowledgement

This research is supported by funding from the Chan-Zuckerberg Initiative.

## Supplementary Table and Figures

**Supp Table 1:** Top 10 sites by the total number of claims across all years for the top 4 medical AI procedures. Each cell represents the major city and state for a unique zip code associated with a medical AI procedure.

| **Coronary Artery Disease** | **Diabetic Retinopathy** | **Liver MR** | **Coronary Atherosclerosis** |
| --- | --- | --- | --- |
| Baton Rouge, LA | Philadelphia, PA | San Francisco, CA | Beverly Hills, CA |
| Royal Oak, MI | Dayton, OH | Philadelphia, PA | Pensacola, FL |
| Pittsburgh, PA | New Orleans, LA | Morristown, NJ | Hiawatha, IA |
| Green Bay, WI | Howell, NJ | Pasadena, CA | Torrance, CA |
| Chattanooga, TN | Santa Fe Springs, CA | San Diego, CA | Eden Prairie, MN |
| Mobile, AL | Green Bay, WI | Burlington, MA | Ann Arbor, MI |
| Evanston, IL | Stanford, CA | Seattle, WA | Dearborn, MI |
| Stanford, CA | Los Angeles, CA | Ann Arbor, MI | Los Angeles, CA |
| Houston, TX | Schertz, TX | Renton, WA | Chicago, IL |
| Atlanta, GA | Minneapolis, MN | Stanford, CA | Troy, NY |

**Supp Figure 1:**
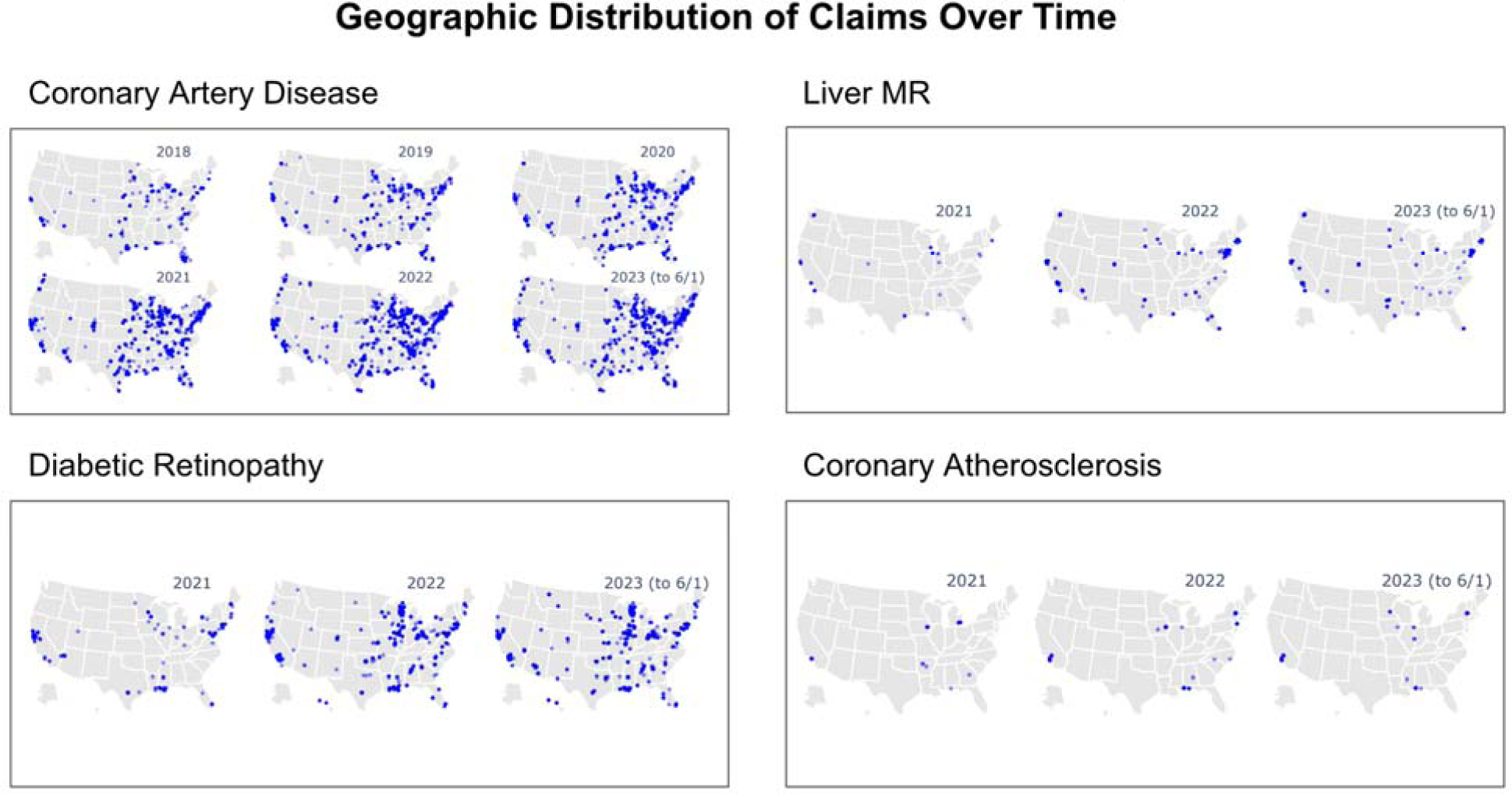
Geographic Distribution of Claims Over Time. The top four most used medical AI procedures are shown geographically across time. Plots for 2023 are recent to 6/1/2023. Each blue dot represents a unique zip code where the AI device is used in a given year. Plots include years from when a medical AI procedure for a device was reported until 2023.

